# Spinal cord damage in Friedreich’s ataxia: Results from the ENIGMA-Ataxia

**DOI:** 10.1101/2022.04.20.22273878

**Authors:** Thiago JR Rezende, Isaac M Adanyeguh, Filippo Arrigoni, Benjamin Bender, Fernando Cendes, Louise A Corben, Andreas Deistung, Martin Delatycki, Imis Dogan, Gary F Egan, Sophia L Göricke, Nellie Georgiou-Karistianis, Pierre-Gilles Henry, Diane Hutter, Neda Jahanshad, James M Joers, Christophe Lenglet, Tobias Lindig, Alberto RM Martinez, Andrea Martinuzzi, Gabriella Paparella, Denis Peruzzo, Kathrin Reetz, Sandro Romanzetti, Ludger Schöls, Jörg B Schulz, Matthis Synofzik, Sophia I Thomopoulos, Paul M Thompson, Dagmar Timmann, Ian H Harding, Marcondes C. França

## Abstract

**Objective:** Spinal cord damage is a hallmark of Friedreich ataxia (FRDA), but its progression and clinical correlates remain unclear. Here we performed a characterization of cervical spinal cord structural abnormalities in a large multisite FRDA cohort.

**Methods:** We performed a cross-sectional analysis of cervical spinal cord (C1 to C4) cross-sectional area (CSA) and eccentricity using MRI data from eight sites within the ENIGMA-Ataxia initiative, including 256 individuals with FRDA and 223 age- and sex-matched controls. Correlations and subgroup analyses within the FRDA cohort were undertaken based on disease duration, ataxia severity, and onset age.

**Results:** Individuals with FRDA, relative to controls, had significantly reduced CSA at all examined levels, with large effect sizes (*d*>2.1) and significant correlations with disease severity (*r*<-0.4). Similarly, we found significantly increased eccentricity (*d*>1.2), but without significant clinical correlations. Subgroup analyses showed that CSA and eccentricity are abnormal at all disease stages. However, while CSA appears to decrease progressively, eccentricity remains stable over time.

**Interpretation:** Previous research has shown that increased eccentricity reflects dorsal column (DC) damage, while decreased CSA reflects either DC or corticospinal tract (CST) damage or both. Hence, our data support the hypothesis that damage to DC and CST follow distinct courses in FRDA: developmental abnormalities likely define the DC, whereas CST alterations may be both developmental and degenerative. These results provide new insights about FRDA pathogenesis and indicate that CSA of the cervical spinal cord should be investigated further as a potential biomarker of disease progression.

## Introduction

Friedreich ataxia (FRDA) is a neurogenetic disease caused by GAA expansions or point mutations in the first intron of the *FXN* gene^1^, leading to lower levels of the protein frataxin and resulting in mitochondrial dysfunction and neurodegeneration^2^. FRDA is the most common autosomal recessive ataxia worldwide^2^. The first symptoms typically begin in late childhood or adolescence and are characterized by slowly progressive ataxia and sensory abnormalities^2,3^. A smaller subset of individuals manifest symptoms after the age of 25 years and are known as people with Late-Onset Friedreich Ataxia (LOFA)^4,5^. These individuals are clinically characterized by slower disease progression and milder non-neurological symptoms^6^.

Pathology studies in FRDA show that structural damage affects both the central and peripheral nervous system^7,8^. In fact, the spinal cord, dorsal root ganglia and dentate nucleus of the cerebellum are the main targets of damage in the disease^7^. MRI-based studies have confirmed such findings and, beyond that, have shown structural damage in the cerebellum, brainstem, cerebellar peduncles and motor cortex^9^.

In recent years, there has been renewed interest in assessing spinal cord damage using non-invasive MRI in FRDA^10-14^. Quantitative structural neuroimaging studies have revealed atrophy and antero-posterior flattening in affected subjects, particularly at cervical and thoracic levels^10,11,14^. Using diffusion tensor imaging (DTI), Hernandez et al (2021) and Joers et al (2022) also reported microstructural changes in the corticospinal tracts and dorsal columns of the cervical spinal cord in individuals with FRDA. Using magnetic resonance spectroscopy, Joers et al (2022) reported large neurochemical changes in the spinal cord in FRDA. In all these studies, the authors were able to find significant correlations between spinal cord MRI metrics and disease severity. Thus, *in vivo* imaging is well-aligned with histological evidence that spinal cord compromise plays a major role in the pathophysiology of FRDA.

Several aspects of spinal cord changes in people with FRDA remain unclear. It is not yet established how spinal cord morphometric abnormalities – atrophy and flattening – change along the disease course. Moreover, differences may exist in the magnitude, progression, and association with clinical variables of spinal cord damage in pediatric *vs* adult patients, and in individuals with early *vs* late symptom onset. These are relevant issues, not only to understand the underlying biology of the disorder, but also to uncover potential imaging-based biomarkers.

Prior neuroimaging studies have generally relied on modest sample sizes from single sites, limiting opportunities to provide robust disease characterization, reliable effect size estimates, and subgroup analyses. The ENIGMA-Ataxia working group is an international collaboration that aggregates MRI data from individuals with ataxias. This consortium offers a unique opportunity to enlarge cohort sizes and to accomplish more detailed analyses in rare diseases, such as FRDA^15^. Hence, the main goal of the present study was to perform a comprehensive evaluation of cervical spinal cord damage in FRDA using a large dataset collected within the ENIGMA-Ataxia group. We sought to characterize the pattern of damage and how it evolves across disease subgroups, stratified according to the time from onset and the magnitude of disease severity.

## Methods

### Participants and Data

We performed a retrospective cross-sectional analysis of data from eight sites in the ENIGMA-Ataxia working group, totaling 256 patients with molecular confirmation of FRDA and 223 age- and sex-matched non-ataxic controls (Table 1, Supplementary Table 1). Disease duration and age at symptom onset were recorded for all participants with FRDA, and disease severity was quantified using one of the following validated clinical scales: the Friedreich Ataxia Rating Scale (FARS)^16,17^, the modified FARS (mFARS)^18^ or the Scale for Assessment and Rating of Ataxia (SARA)^19^. To assess the cervical spinal cord, we used high-resolution T1-weighted MRIs covering the brain and upper cervical vertebrae acquired on 3T clinical scanners with spatial resolution not inferior to 1-mm isotropic (Supplementary Table 2). Individuals with FRDA and controls from each site underwent MRI scans using the same scanner and protocol.

**Table 1:** Demographics data for all sites. For the healthy controls demographics data, please see Supplementary Table 1.

| Sites | Age (years) | Sex |  | GAA1 | GAA2 | Onset Age (years) | Disease Duration (years) | Disease Severity |  |  |
| --- | --- | --- | --- | --- | --- | --- | --- | --- | --- | --- |
|  | Average [Range] | Male | Female | Average | Average | Average | Average | Scale | Average | Normalized Scale |
| **Aachen (N=32)** | 36±12 [19-59] | 16 | 16 | 497±224 | 819±218 | 17±8 | 20±9 | SARA | 20±9 | 0.486±0.227 |
| **Campinas (N=83)** | 30±13 [7-66] | 31 | 52 | 1026±267 | 869±215 | 18±9 | 12±10 | FARS | 55±22 | 0.440±0.178 |
| **Conegliano (N=46)** | 24±12 [8-51] | 18 | 21 | 671±179 | - | 12±7 | 13±9 | SARA | 18±8 | 0.441±0.199 |
| **Essen (N=15)** | 44±11 [26-60] | 6 | 9 | 415±292 | 648±306 | 17±10 | 23±9 | SARA | 24±4 | 0.598±0.107 |
| **Melbourne1 (N=22)** | 39±14 [22-63] | 11 | 11 | 532±234 | 908±222 | 21±9 | 18±10 | FARS | 84±29 | 0.667±0.226 |
| **Melbourne2 (N=14)** | 30±9 [18-49] | 8 | 6 | 604±206 | 870±186 | 15±4 | 14±7 | mFARS | 47±22 | 0.526±0.164 |
| **Minnesota (N=26)** | 19±7 [10-35] | 14 | 12 | 598±184 | 960±213 | 14±5 | 6±4 | FARS* | 43±14 | 0.341±0.111 |
| **Tubingen (N=14)** | 32±11 [18-53] | 10 | 4 | - | - | 18±9 | 14±7 | SARA | 18±8 | 0.447±0.208 |

\*Maximum score 117.

Data collection, analysis, and contributions to this project were approved by the human research ethics body at each site as defined here: Aachen (IRB RWTH Aachen University, project EK 083/15); Campinas (IRB University of Campinas, project CAAE 29869520.8.3001.5404); Conegliano (IRB IRCCS Eugenio Medea, project 155/CE-Medea); Essen (IRB Essen University Hospital, project 15-6404-BO); Melbourne (IRB Monash Health, project #13201B)^15^; Minnesota (IRB University of Minnesota, STUDY00009047); Tubingen (IRB University of Tuebingen, project 598/2011BO1). Multisite data aggregation and analysis was approved the Monash University Human Research and Ethics Committee (project #12372). All data was fully anonymized prior to aggregation, including assignment of new subject identifier codes.

### Image Processing

Data processing was undertaken using harmonized protocols developed by the ENIGMA-Ataxia consortium (http://enigma.ini.usc.edu/ongoing/enigma-ataxia/), based on publicly available and well-validated software toolboxes^20^.

To measure the cross-sectional area (CSA) and eccentricity, we employed the Spinal Cord Toolbox (SCT) version 4.2.2, an open-source software package specifically designed to process spinal cord multimodal MRI data^20^. In brief, automatic segmentation of the cervical spinal cord was conducted using a deep-learning algorithm^21^ and, if deemed necessary after visual inspection, the segmentations were manually corrected. Next, the C2 and C3 vertebral levels were manually marked at the posterior tip of the vertebral discs, which enabled the registration of subject images to a standardized template of the spinal cord and brainstem (the PAM50 template)^22-24^. Lastly, the mean CSA and eccentricity were computed at each of the C1 to C4 vertebrae after correcting for the curvature of the spine. The CSA is quantified by the number of pixels in the set of axial slices defining each vertebral level of the segmented spinal cord, reported in millimeters squared. Eccentricity is computed by fitting an ellipse to each axial spinal slice and determining the deviation (i.e., flattening) of the ellipse relative to a perfect circle. Mathematically, such a measure characterizes the shape of the spinal cord cross-section defined as the square root of 1 - (*d*/*D*)^2^, where *d* and *D* are respectively defined as the smallest and largest diameter of the ellipse. Values closer to 1 indicate an antero-posterior flattening of the spinal cord. We only assessed the upper cervical spinal cord, since we used MR images centered on the brain with limited spinal cord coverage (Figure 1). Since the spinal cord coverage was slightly different across individuals due to head size variability or field-of-view placement during data acquisition, different sample sizes were available for each vertebral level we examined (Controls: C1=223, C2=223, C3=215 and C4=170; Patients: C1=252, C2=252, C3=237 and C4=170).

**Figure 1:**
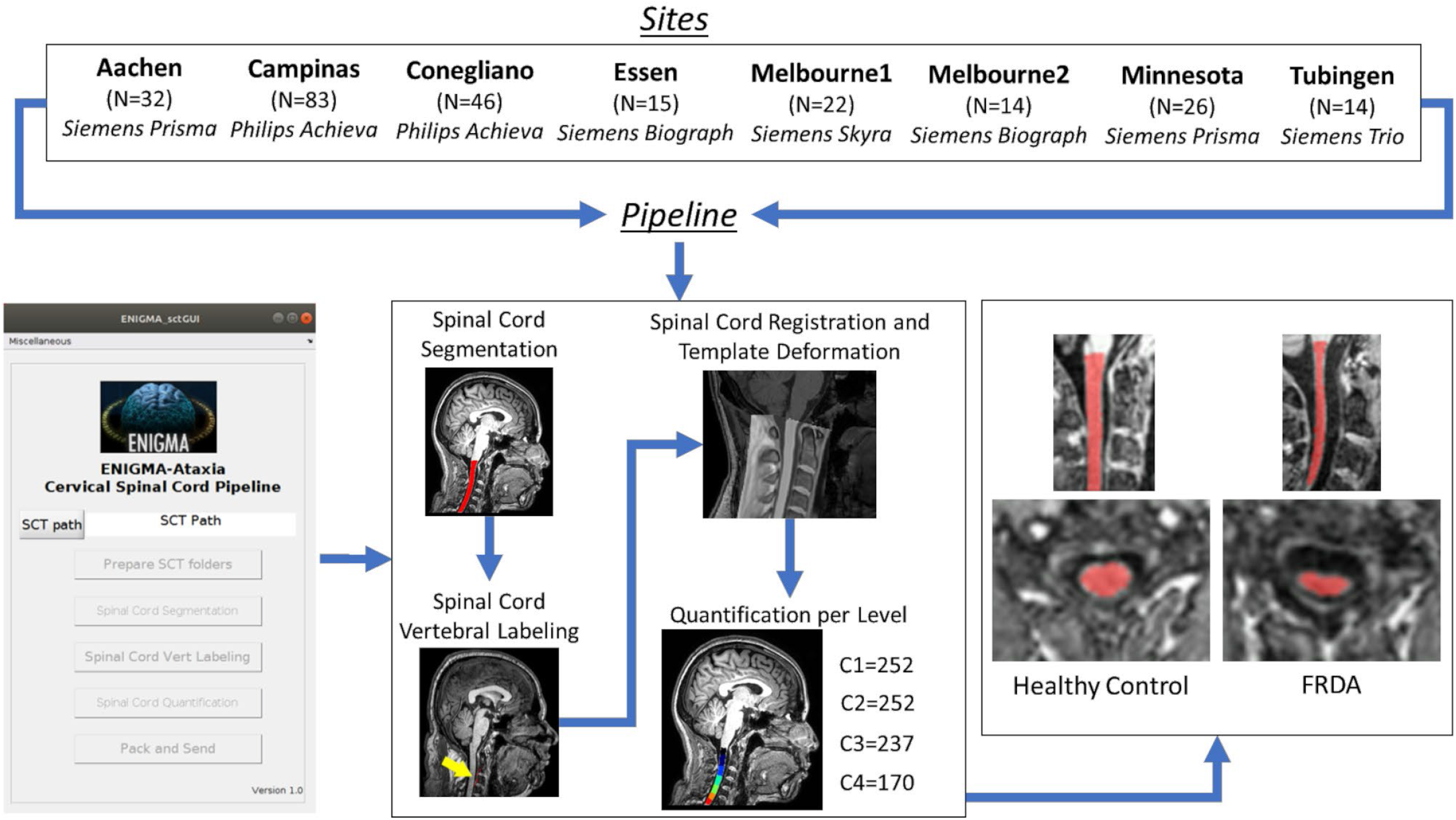
Study design and imaging processing pipeline. For healthy controls numbers, please see Supplementary Table 1.

### Statistical Analysis

#### Overall FRDA vs. Control Comparison

We compared CSA and eccentricity at each vertebral level from C1 to C4 in all individuals with FRDA relative to the age- and sex-matched control cohort using ANCOVAs with age, sex and site as covariates of no interest. We corrected for multiple comparisons using Bonferroni adjustment of statistical significance thresholds. Effect sizes (ES) of statistically significant results were computed as follows (Cohen’s *d*):

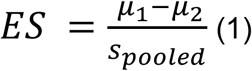

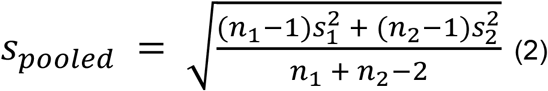

where, μ_1_ and μ_2_ are the mean values for the control and FRDA groups respectively, s_pooled_ is the pooled standard deviation, n_1_ and n_2_ are the number of subjects in each group, and s_1_ and s_2_ are the respective group standard deviations. We considered effect size values of 0.2 as small, 0.5 as moderate, 0.8 as large, and > 1.2 as very large, according to established convention^25^.

#### Correlation Analysis

To assess correlations between spinal cord morphometric data (CSA and eccentricity) and clinical parameters (disease duration and disease severity), we used the Pearson correlation coefficient. Before performing the analyses, we first adjusted the data to account for site, age and sex effects using a linear model. Multiple clinical rating scales were used to assess disease severity across the sites (FARS, mFARS and SARA). There was a high correlation between SARA and FARS total neurological scores (r=0.860 and p<0.0001) in our participants for whom both scales were collected at the same time, which is in agreement with comparable previous work from Bürk and colleagues (2009; r=0.953 and p<0.0001)^26^. To accomplish a direct pooled analysis, we therefore created a normalized disease severity variable by dividing the disease severity scores by the respective maximum value of the respective scale, e.g., disease severity measures quantified using FARS were divided by 125 (except by Minnesota site, max score 117), mFARS were divided by 93 and SARA measures were divided by 40. We note that these scales likely have slightly different psychometric properties (e.g., differing ceiling and floor effects)^18,27^, and thus while this normalization approach is strongly supported by the very high inter-scale correlations, we acknowledge that there is not a precise 1-to-1 correspondence in their absolute or relative scores. However, our goal is not to establish absolute harmonization across scales or investigate detailed symptom expression and progression, but rather to test for general trends between overall ataxia severity and spinal cord structure.

#### Clinical Subtype Comparison

We also analyzed spinal cord damage in specific clinical subgroups: pediatric patients (age <18 years) and individuals with LOFA (first onset of symptoms at age >25 years). Subgroups of controls were selected to match each clinical subgroup for age, sex, and site. The pediatric group included 40 individuals aged <18 years (Controls: N=26, mean age=14.0±2.4, 12M/14F; FRDA: N=40, mean age=13.3±2.5, 18M/22F), and the second group 45 individuals with LOFA (Control: N=42 mean age=45.4±9.3, 22M/20F; LOFA: N=45, mean age=45.6±8.8, 25M/20F). Between-group comparisons of spinal cord measures (CSA and eccentricity) and correlations with clinical parameters (disease duration and severity) were undertaken in each subgroup as described above.

#### Disease Progression

To examine spinal cord differences across different disease stages, we defined five subgroups (DD1 – DD5) according to disease duration (time since first symptom expression) at the time of each participant’s scan: <5 years, 5-10 years, 11-15 years, 16-20 years, and >20 years respectively. For further characterization of the data, we also defined four subgroups (DS1 – DS4) according to the normalized disease severity of each participant’s scan: <0.25, 0.26-0.50, 0.51-0.75 and >0.75 respectively. These divisions do not represent clinically-determined cut-offs, but rather provide an intuitive means of quantitatively assessing and reporting changes in effect sizes with disease progression. We first compared each subgroup with a non-ataxic control cohort matched by age, sex and site. Subsequently, we compared each subgroup with the earliest (DD1) or least severe (DS1) subgroup to assess evidence for progressive degeneration independent of early/pre-symptomatic effects. Similar to the statistical approach used in the general comparison, we used ANCOVA to assess each group’s differences, using age, sex and site as covariates and used Bonferroni correction to adjust for multiple comparisons.

## Results

### Overall FRDA vs Control Comparison

Individuals with FRDA relative to controls had significantly reduced CSA at all vertebral levels (Figure 2a) with very large effect sizes (C1 ES=2.6, C2 ES=2.6, C3 ES=2.3, C4 ES=2.1). Similarly, we found significantly increased eccentricity at all vertebral levels (Figure 2b), also with very large effect sizes (C1 ES=1.2, C2 ES=1.4, C3 ES=1.3, C4 ES=1.4), although substantially smaller in comparison to CSA. In addition, the spinal cord growth curve, i.e., the plot of spinal cord CSA *vs* age, revealed distinct patterns in the control group (C1: r=-0.050, p=0.999; C2: r=-0.045, p=0.999; C3: r=-0.068, p=0.999; C4: r=-0.039, p=0.999), CSA remains stable over the entire lifespan, whereas in individuals with FRDA (C1: r=-0.247, p<0.001; C2: r=-0.216, p=0.003; C3: r=-0.227, p=0.002; C4: r=-0.244, p=0.006), CSA appears to show a progressive decline with age (Figure 3).

**Figure 2:**
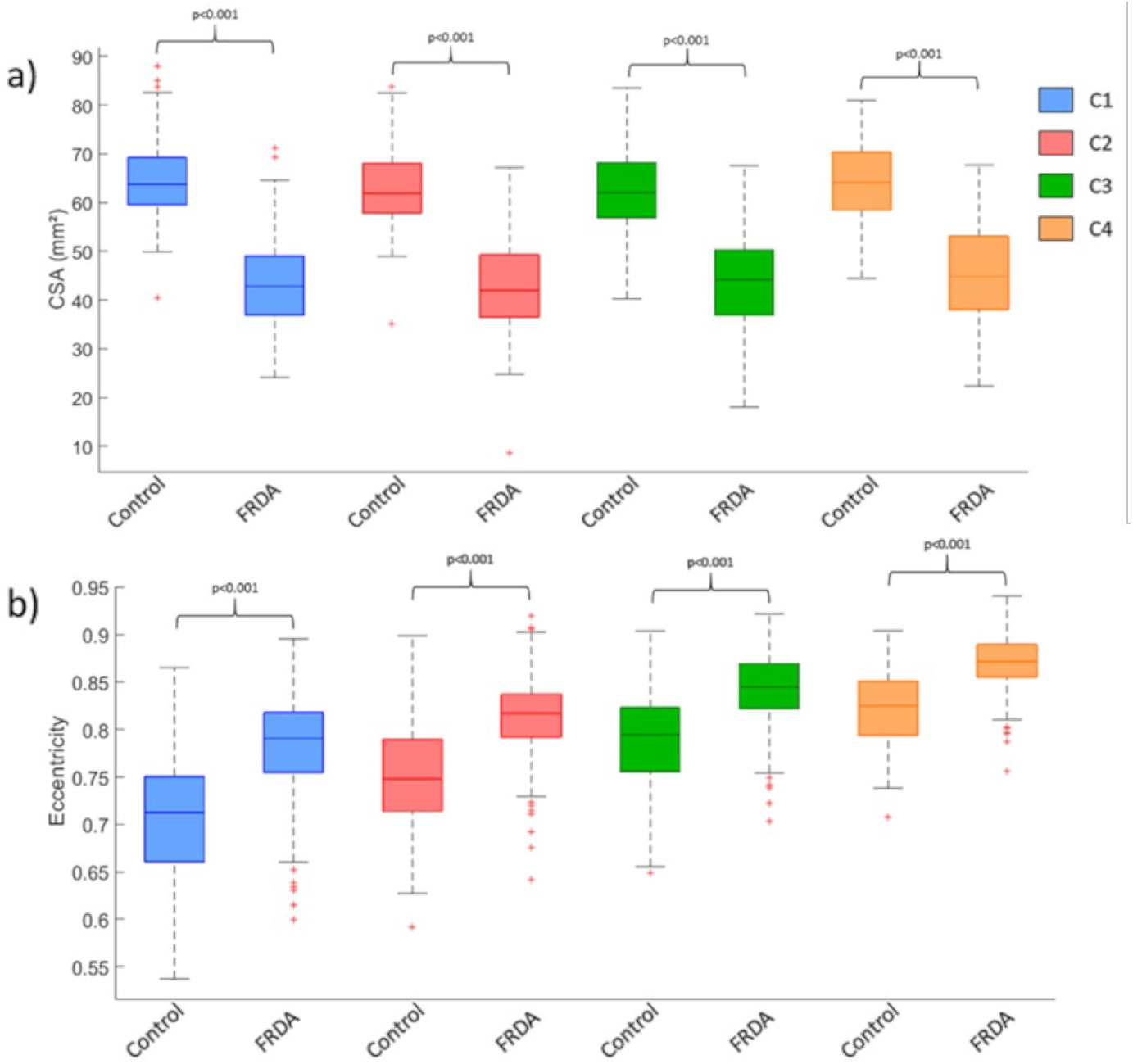
Box plots displaying group differences at each spinal cord segment, C1-C4, for the total cohort. a) Cross-sectional area in square millimeters; b) Eccentricity.

**Figure 3:**
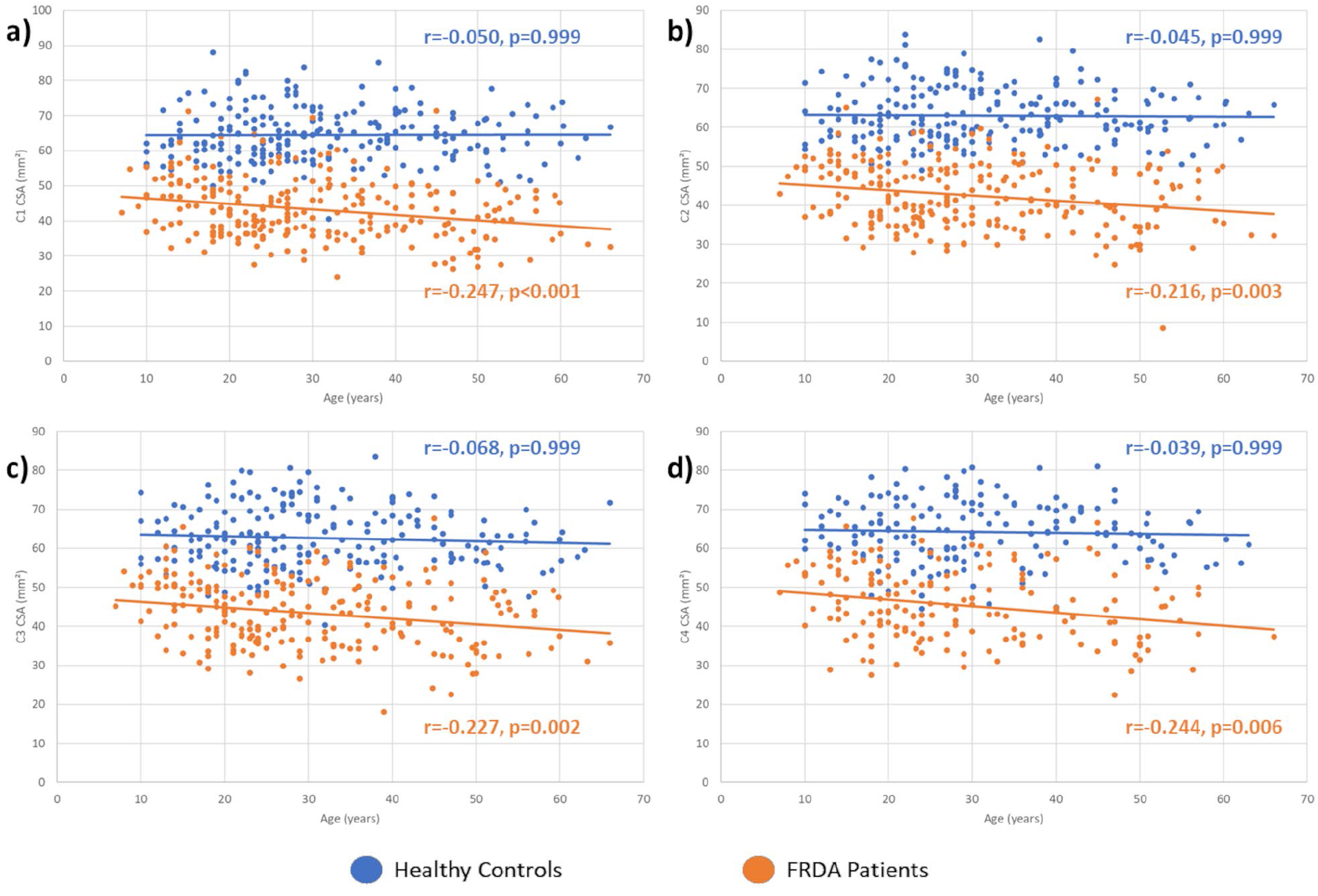
Plot of spinal cord cross-sectional (CSA) *versus* age in patients and controls for vertebral level a) C1, b) C2, c) C3 and d) C4.

### Correlation Analysis

We found significant correlations between the normalized disease severity or ataxia duration and CSA at all vertebral levels assessed (C1-C4) (Figure 4) after Bonferroni adjustment for multiple comparisons (Normalized disease severity - C1: r=-0.424, p<0.001; C2: r=-0.395, p<0.001; C3: r=-0.399, p<0.001; C4: r=-0.435, p<0.001. Ataxia Severity - C1: r=-0.174, p=0.006; C2: r=-0.146, p=0.044; C3: r=-0.164, p=0.026; C4: r=-0.237, p=0.004). In contrast, we did not find any significant correlation between eccentricity and normalized disease severity or ataxia duration.

**Figure 4:**
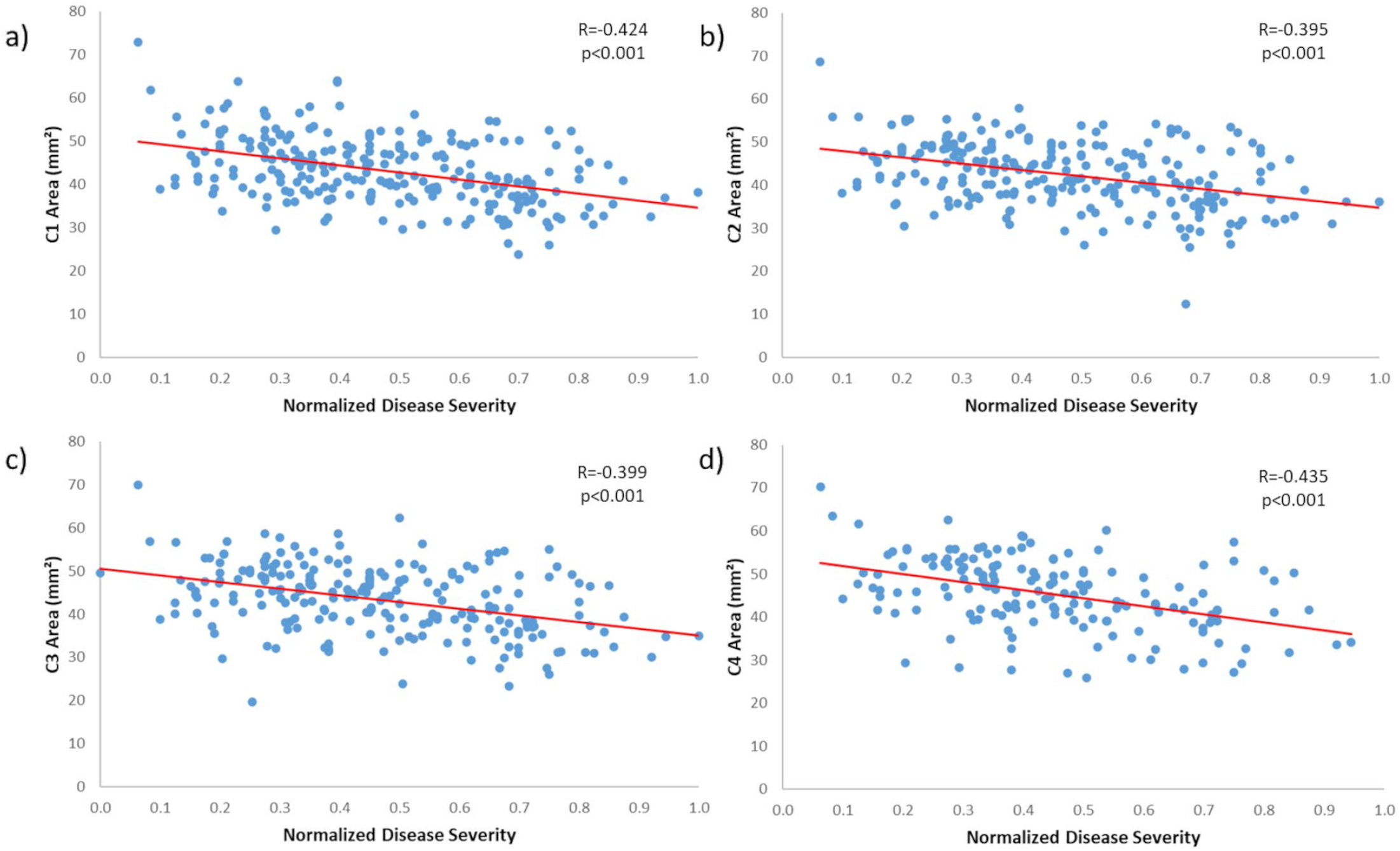
Significant correlations between normalized disease severity and cross-sectional area in individuals with FRDA at vertebral level a) C1, b) C2, c) C3 and d) C4.

### Comparison of Clinical Subtypes

Children with FRDA showed abnormal CSA and eccentricity when compared to matched non-ataxic controls (Figure 5a) with very large effect sizes (CSA: C1 ES=1.7, C2 ES=2.1, C3 ES=2.0, C4 ES=2.1; eccentricity: C1 ES=1.3, C2 ES=1.8, C3 ES=1.8, C4 ES=1.5). Differences relative to adults with FRDA (with ‘classical’ onset age) matched by age-sex or disease duration did not reach statistical significance (Supplementary Table 3).

**Figure 5:**
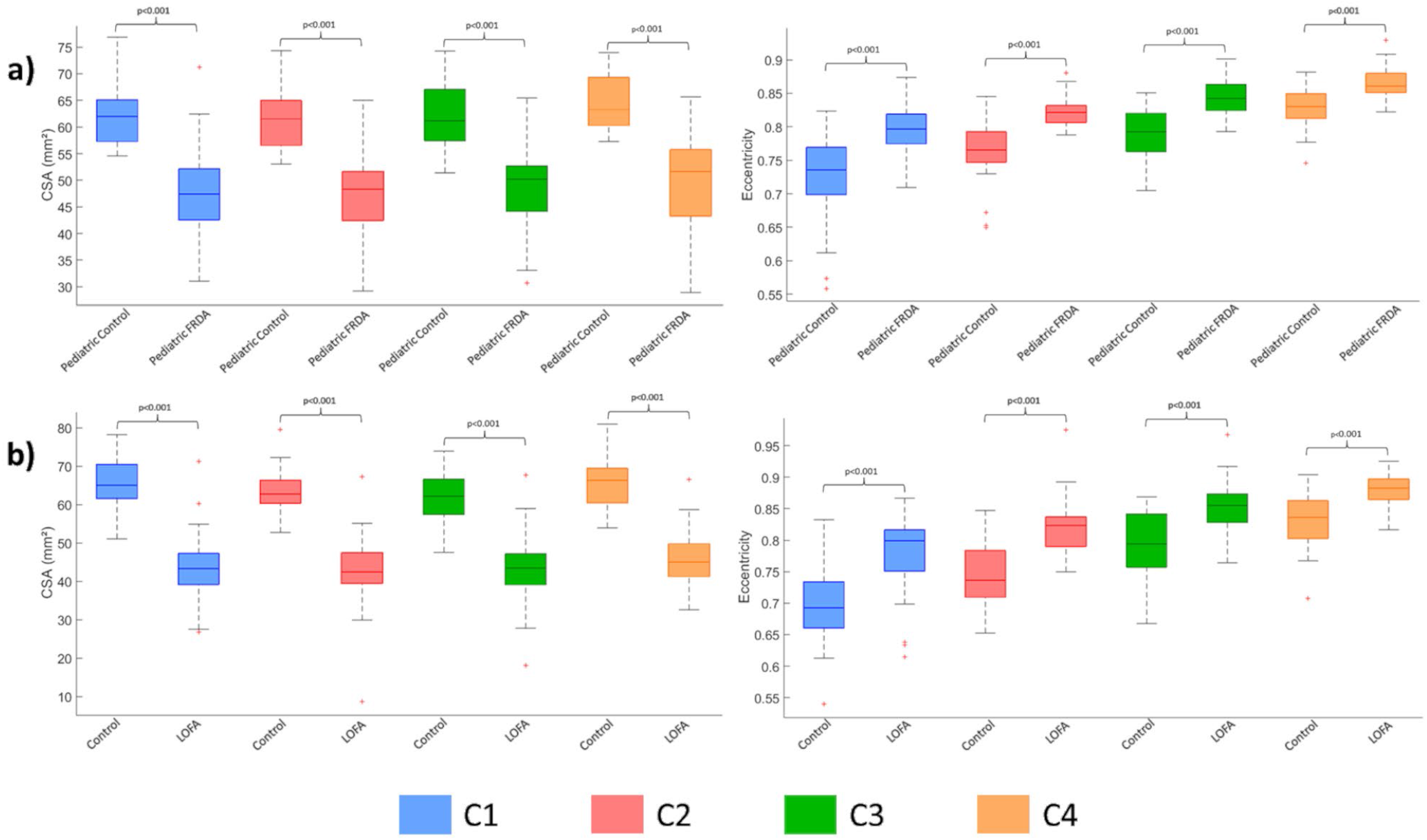
Box plots displaying group differences at each spinal cord level, C1-C4 a) children (age <18 years) with FRDA vs matched controls; and b) individuals with late-onset Friedreich ataxia (LOFA) vs matched controls.

Individuals with LOFA showed a similar result when compared to matched non-ataxic controls (Figure 5b) (CSA: C1 ES=3.0, C2 ES=2.9, C3 ES=2.6, C4 ES=2.3; eccentricity: C1 ES=1.4, C2 ES=1.7, C3 ES=1.4, C4 ES=1.2). Similarly, differences relative to adults with classical FRDA matched by age-sex or disease duration did not show statistical significance.

We did not find significant correlations between spinal cord measures and clinical variables in the pediatric cohort (Supplementary Table 4). However, for individuals with LOFA, we found significant correlations between CSA and normalized disease severity for all vertebral levels assessed, except for C4, after Bonferroni correction (C1: r=-0.385, p<0.010; C2: r=-0.371, p=0.026; C3: r=-0.357, p=0.035).

### Disease Evolution

#### Disease Duration

The subgroup analyses based on disease duration showed that CSA and eccentricity are already abnormal in the earliest stages of the disease, with significant differences relative to controls in all subgroups (Figures 6 and 7). In addition, we found significantly reduced CSA when DD3 (10-15yrs post-symptom duration), DD4 (15-20yrs duration) and DD5 (20+ years duration) were compared to DD1 (<5 years duration) at all vertebral levels (Figure 5 and 6). In contrast, eccentricity remained stable across the subgroups.

**Figure 6:**
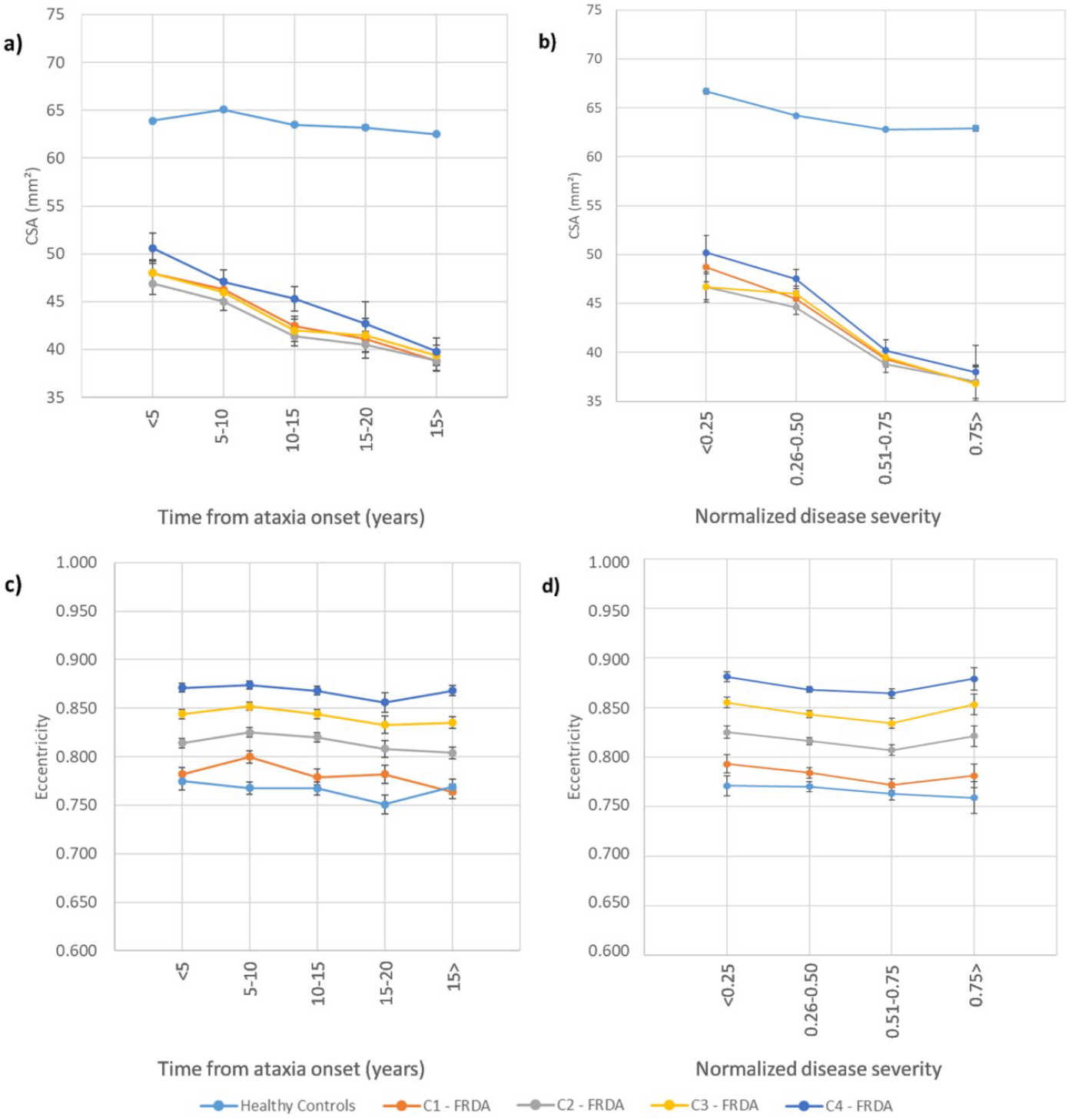
Results showing the progressive atrophy of the cervical spinal cord area (CSA) (a, b) and eccentricity (c, d) in participants with FRDA and healthy controls. Panels a) and c) depict subgroups based on disease duration (DD); b) and d) show subgroups based on disease severity (DS). To the healthy controls, the measures represent the mean cervical spinal cord area or eccentricity; error bars = standard error of the mean. Subgroups based on disease duration, DD1: Time from ataxia onset <5 years, DD2: Time from ataxia onset between 5-10 years, DD3: Time from ataxia onset between 10-15 years, DD4: Time from ataxia onset between 15-20 years, DD5: Time from ataxia onset >20 years. Subgroups based on disease severity, DS1: Normalized disease severity <0.25, DS2: Normalized disease severity between 0.26-0.50, DS3: Normalized disease severity between 0.51-0.75, DS4: Normalized disease severity >0.75.

**Figure 7:**
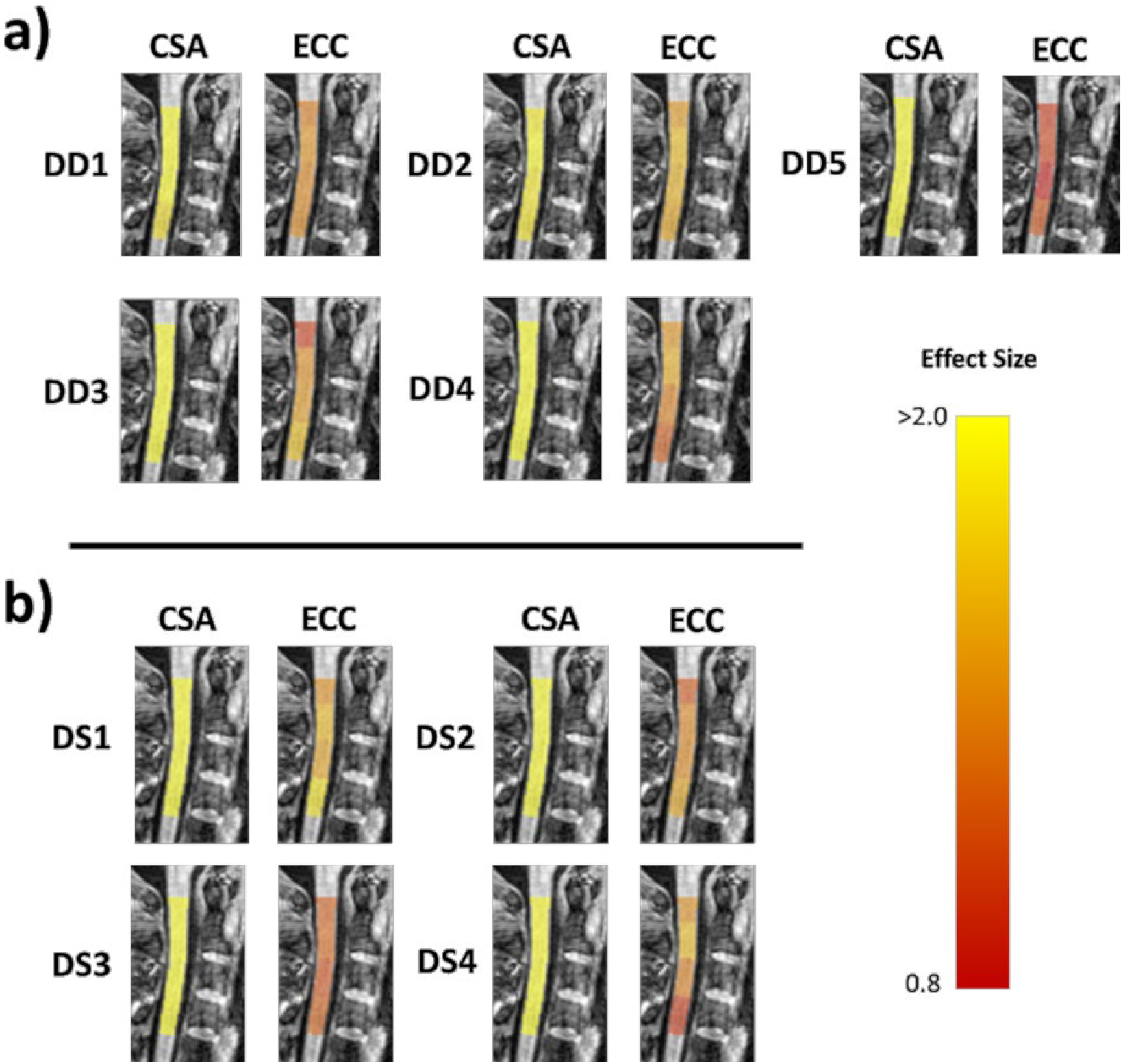
Effect size values for CSA and eccentricity at each stage of FRDA. a) Subgroups based on disease duration, DD1: Time from ataxia onset <5 years, DD2: Time from ataxia onset between 5-10 years, DD3: Time from ataxia onset between 10-15 years, DD4: Time from ataxia onset between 15-20 years, DD5: Time from ataxia onset >20 years. b) Subgroups based on disease severity, DS1: Normalized disease severity <0.25, DS2: Normalized disease severity between 0.26-0.50, DS3: Normalized disease severity between 0.51-0.75, DS4: Normalized disease severity >0.75.

#### Disease Severity

The subgroup analyses based on disease severity showed similar results. Abnormalities in CSA and eccentricity are observable in patients with normalized disease severity <0.25, with significant effects relative to controls in all subgroups (Figure 6 and 7). We also found significantly reduced CSA when DS3 (normalized disease severity 0.51-0.75) and DS4 (severity >0.75) were compared to DS1 (severity <0.25) at all vertebral levels. Meanwhile, DS2 (severity 0.26-0.50) showed reduced CSA, relative to DS1, only for C1 and C2; eccentricity remained stable across the subgroups.

## Discussion

Spinal cord damage has been recognized as a hallmark of FRDA since Nikolaus Friedreich’s first reports and confirmed in more recent histology and neuroimaging studies^7,10-14,28^. In this study, we performed a retrospective cross-sectional analysis of cervical spinal cord structure using MRI data from a large multisite cohort. We report significant and substantial CSA reduction in individuals with FRDA at all vertebral levels examined, relative to non-ataxic individuals, and significant correlations with disease severity scores. Eccentricity differences were also pronounced in this cohort relative to controls, but effect sizes were smaller than for CSA and no significant clinical correlations were observed. Subgroup analyses based on disease duration and severity showed that CSA and eccentricity are already abnormal in the early stages of the disease and that CSA likely declines with disease progression, whereas eccentricity remains stable. Taken together, CSA emerges as a potential MRI biomarker candidate for clinical tracking in FRDA.

Our results are consistent with previous MRI-based studies that found cervical spinal cord atrophy and anteroposterior flattening in FRDA^10-14,29^. *Post-mortem* studies indicate that the pathological correlates of these findings are severe depletion of myelinated fibers in the dorsal columns, dorsal spinocerebellar and lateral corticospinal tracts^28^. These findings are also consistent with a single-site prospective study that showed a decrease in CSA over time in individuals with FRDA in an early-stage cohort, with no decrease over time in eccentricity^14^.

Prior studies undertaken in other spinal cord diseases help us understand the pathological underpinnings of changes in CSA and eccentricity^30-33^. Indeed, different patterns emerge when one compares diseases characterized by predominant/exclusive lateral column involvement (e.g., amyotrophic lateral sclerosis, pure subtypes of hereditary spastic paraplegia) *vs* diseases with predominant/exclusive dorsal column involvement (e.g., acquired sensory neuronopathies)^30-32^. CSA reduction is evident in both groups, but eccentricity increase is only reported in the latter^30^. Therefore, eccentricity can be considered a surrogate MRI marker for dorsal column damage, whereas CSA may be related to abnormal integrity in both lateral and dorsal columns. Using this conceptual framework, relevant insights can be inferred from our results. The stability of eccentricity alongside decreasing CSA across FRDA stages (based on duration or severity) suggests that the corticospinal tract and dorsal columns follow distinct mechanisms and time courses of damage in the disease. Corticospinal tract damage is most consistent with a combination of abnormal developmental and progressive degenerative processes, as shown by both early (already seen in the pediatric subgroup) and progressive CSA abnormalities. In contrast, dorsal column abnormalities, assessed by eccentricity, may be related to early maldevelopment but remain stable along the entire disease course, at least from the point of first symptom expression.

Our assumption that dorsal column damage is neurodevelopmental is in line with neuropathological reports from Koeppen et al (2017). These authors suggest that the developmental failure of the dorsal root ganglia (DRG) leads to the secondary hypoplasia of dorsal columns, since DRG are the source of myelinated fibers in the dorsal columns. Indeed, the autopsy of two young patients with FRDA showed that the neurons in the dorsal nuclei were severely reduced or absent, probably due to the lack of innervation from the dorsal root collaterals that occurs during the gestational period^34^, arguing in favor of a developmental failure. Furthermore, experiments using animal models provide evidence that frataxin plays a role in embryonic development^34,35^. Our imaging data suggest that CSA of individuals with FRDA, on average, reaches its maximum before 10 years of age and then starts to decrease, whereas healthy controls have higher CSA values relative to FRDA patients even at the earliest disease stages, and keep stable over time. A preceding MRI-based study performed by Rezende and colleagues (2019)^12^ found a very similar result.

Progressive neurodegeneration in the corticospinal tract is consistent with the hypothesis that pyramidal tract damage in FRDA arises from a ‘dying back’ process. Neuropathological studies have found that the corticospinal tract is more affected in the spinal cord than in the brain, with the exception of the lack of Betz cells in the motor cortex^36,37^. Koeppen and Mazurkiewicz (2013) also showed that spinal cord damage is more severe in thoracic levels compared to cervical regions. Previous neuroimaging studies also support such a concept. Indeed, a diffusion MRI-based study showed that microstructural abnormalities were more robust in caudal levels, although clinical correlations were stronger at the upper levels of the corticospinal tract (Hernandez et al, 2021). Rezende and colleagues (2019) also reported motor cortex thinning only in adults, but not in children with FRDA, alongside progressive damage in the cerebral corticospinal tract. This is in agreement with Harding and colleagues (2021) who proposed a disease staging schema for brain damage in FRDA. These authors highlight the progressive pattern of damage in the disease that begins in infratentorial structures and spreads to cortical structures in later disease stages^15^.

From a clinical perspective, our data indicate that CSA at C1 level is a potential biomarker candidate as it showed the highest correlation coefficient with disease severity and the highest effect size compared to controls. CSA at C1 level also had the highest effect size in a recent single-site longitudinal study^14^. However, this may not be the case for all FRDA stages or sub-phenotypes. For the pediatric cohort (age <18 years), we did not find any significant correlations between CSA and normalized disease severity, whereas such associations were evident in the adult cohort. Although this observation may reflect statistical power as there were fewer pediatric individuals (n=40) relative to adults (n=159), a similar result was previously reported by Rezende and colleagues (2019). In a recent paper, Hernandez et al. showed higher correlation coefficients between diffusion MRI-based parameters of the corticospinal tract and disease severity for C2 and C3 respectively. Therefore, we propose that the ideal neuroimaging biomarker may vary according to the cohort profile under evaluation. There might exist a specific neuroimaging biomarker for each disease stage, similar to what has been suggested for SCA3^38^. This hypothesis is supported by the proposed mechanism of corticospinal degeneration in FRDA, which seems to follow a dying-back motor axonopathy. Lower levels of the spinal cord may therefore already be extensively impacted very early in the disease course and reach an early floor effect. Ongoing damage to the spinal corticospinal tract may therefore be more easily captured by MRI metrics at upper levels. At this point, prospective studies with pediatric and adult cohorts must be undertaken to confirm such hypotheses.

Notwithstanding the original contributions of this study, several limitations must be acknowledged. This is a cross-sectional study and many of the findings presented here must be confirmed by prospective longitudinal neuroimaging studies, particularly those enriched with a pediatric cohort. Our analyses were performed using T1-weighted brain MRI, which is the most common and widely used MRI sequence for research and clinical practice. However, this confines our assessment to the upper portions of the cervical spinal cord. More targeted spinal cord imaging acquisitions would enable more detailed analyses, such as tract-specific microstructural evaluation and individual assessment of white matter and grey matter regions. Lastly, the use of different clinical scales at each site limits more extensive investigation of correlations between spinal cord damage and disease severity. Here, we also employ a relatively blunt normalization approach to pool scores across different clinical scales. Future work modeling the relationship between different clinical scales (i.e., SARA and FARS) would be beneficial to establish more specific conversion scores. Prospective natural history imaging studies (e.g., TRACK-FA; https://www.monash.edu/medicine/trackfa) will also be key to addressing many of these limitations.

To conclude, our data support the hypothesis that damage to spinal dorsal column and corticospinal tract follow distinct courses in the disease: developmental damage likely defines the former, whereas alterations in the latter may be both developmental and degenerative in origin. These results provide new insights about FRDA pathogenesis and indicate that spinal cord MRI may be a useful biomarker to track disease progression.

## Data Availability

All code and data processing instructions are available at https://github.com/Harding-Lab/enigma-ataxia

## Acknowledgments

The methods of harmonization and multi-site data analysis elements of this work were supported by NIH Big Data to Knowledge (BD2K) program grant number U54 EB020403, and grants from the Australian National Health and Medical Research Council (Fellowship 1106533, Grant 1184403). FARA (Friedreich’s Ataxia Research Alliance, grant 92133) and FAPESP (São Paulo Research Foundation) also supported this study through CEPID/BRAINN (grant 2013/07559-3), the German Research Foundation (DFG, DE 2516/1-1 and TI 239/17-1) and by the European Joint Programme on Rare Diseases (EJPRD), under the EJP RD COFUND-EJP N° 825575 as part of the PROSPAX consortium (to M.S. and D.T. via DFG, German Research Foundation). CMRR at the University of Minnesota is supported by grants NIH P41 EB027061 and P30 NS076408. PGH and CL also acknowledge support by grants from the FARA, GoFAR, Ataxia UK and the Bob Allison Ataxia Research Center.

## Author Contributions

Conception and design of the study & methods: TJRR, IHH, MCF, NJ, SIT, GFE, PMT.

Acquisition and analysis of data: *Aachen:* KR, ID, SR, JBS; *Campinas*: MCF, TJRR, ARMM, FC; *Conegliano*: AM, FA, MV, DP, ST; *Essen*: DT, AD, SLG; *Melbourne*: NGK, IHH, LAC, MBD, GFE; *Minnesota*: CL, PGH, JJ, IMA, DH; *Tubingen*: BB, TL, MS, LS.

Analysis of multi-site data & drafting initial manuscript: TJRR, IHH, MCF.

