## Supplementary Table1 for "Spinal cord damage in Friedreich’s ataxia: Results from the ENIGMA-Ataxia"

**Supplementary Table1:** Demographics data for non-ataxic individuals for each site.

| Sites | Age (years) | Sex | |
| --- | --- | --- | --- |
|  | **Average [Range]** | **Male** | **Female** |
| Aachen (N=34) | 36±12 [22-63] | 15 | 19 |
| Campinas (N=84) | 30±13 [10-66] | 32 | 52 |
| Conegliano (N=23) | 28±9 [16-47] | 10 | 13 |
| Essen (N=13) | 46±10 [28-60] | 6 | 7 |
| Melbourne1 (N=20) | 42±13 [19-62] | 7 | 13 |
| Melbourne2 (N=15) | 27±6 [19-40] | 9 | 6 |
| Minnesota (N=20) | 20±7 [10-35] | 11 | 9 |
| Tubingen (N=14) | 31±9 [19-49] | 9 | 5 |
