## Supplementary Table2 for "Spinal cord damage in Friedreich’s ataxia: Results from the ENIGMA-Ataxia"

**Supplementary Table2:** Scanner and imaging acquisition details for each site.

|  | **Site** | | | | | | | |
| --- | --- | --- | --- | --- | --- | --- | --- | --- |
|  | **Aachen** | **Campinas** | **Conegliano** | **Essen** | **Melbourne1** | **Melbourne2** | **Minnesota** | **Tubingen** |
| **Scanner** | Siemens Prisma | Philips Achieva | Philips Achieva | Siemens Biograph | Siemens Skyra | Siemens Biograph | Siemens Trio and Prisma | Siemens Trio |
| **Field strength (T)** | 3 | 3 | 3 | 3 | 3 | 3 | 3 | 3 |
| **Head coil (channels)** | 64 | 8 | 32 | 16 | 32 | 32 | 32 (Trio) 64 (Prisma) | 32 |
| **Sequence** | MPRAGE | SPGR | SPGR | MPRAGE | MPRAGE | MP2RAGE | MPRAGE | MPRAGE |
| **TR (ms)** | 2400 | 7 | 8 | 2530 | 1540 | 5000 | 2530 | 2300 |
| **TE (ms)** | 2.36 | 3.201 | 3.5 | 3.26 | 2.55 | 3.43 | 3.65 | 3.51 |
| **TI (ms)** | 1000 | - | - | 1100 | 900 | 700, 2500 | 1100 | 900 |
| **Flip (deg)** | 8 | 8 | 8 | 7 | 9 | 4, 5 | 7 | 9 |
| **Plane** | Sagittal | Sagittal | Sagittal | Sagittal | Sagittal | Sagittal | Coronal | Sagittal |
| **Slices** | 208 | 180 | 160 | 176 | 208 | 192 | 224 | 176 |
| **FOV (mm x mm)** | 288x288 | 240x240 | 256x256 | 256x256 | 256x256 | 256x240 | 256x176 | 256x240 |
| **Voxel Size (mm, x y z)** | 0.8x0.8x0.8 | 1x1x1 | 1x1x1 | 1x1x1 | 1x1x1 | 1x1x1 | 1x1x1 | 1x1x1 |
