## Supplementary Table3 for "Spinal cord damage in Friedreich’s ataxia: Results from the ENIGMA-Ataxia"

**Supplementary Table3:** Group differences between pediatric vs adult patients with FRDA at each spinal cord level, C1-C4.

| Veterbral Labels | Variables | pediatric FRDA (mean±SD) | adult FRDA (mean±SD) | p-value | Effect Size |
| --- | --- | --- | --- | --- | --- |
| C1 | CSA (mm²) | 47.9 ± 8.4 | 42.0 ± 8.5 | 0.107 | 0.69 |
|  | ECC | 0.797 ± 0.035 | 0.778 ± 0.058 | 1.000 | 0.36 |
| C2 | CSA (mm²) | 46.9 ± 7.3 | 41.3 ± 7.9 | 0.035 | 0.72 |
|  | ECC | 0.821 ± 0.022 | 0.811 ± 0.046 | 0.508 | 0.24 |
| C3 | CSA (mm²) | 48.3 ± 7.4 | 42.0 ± 8.6 | 0.072 | 0.76 |
|  | ECC | 0.845 ± 0.025 | 0.840 ± 0.042 | 0.799 | 0.14 |
| C4 | CSA (mm²) | 49.3 ± 8.5 | 43.9 ± 9.5 | 0.826 | 0.58 |
|  | ECC | 0.866 ± 0.024 | 0.867 ± 0.032 | 1.000 | 0.06 |
