## Supplementary Table4 for "Spinal cord damage in Friedreich’s ataxia: Results from the ENIGMA-Ataxia"

**Supplementary Table4:** Correlations between time from ataxia onset or normalized disease severity and CSA or eccentricity for the pediatric cohort.

| Time from ataxia onset (years) | | | | |
| --- | --- | --- | --- | --- |
|  | **CSA (mm²)** | | **Eccentricity** | |
|  | **r** | **p-value** | **r** | **p-value** |
| C1 | 0.075 | 0.999 | 0.293 | 0.133 |
| C2 | 0.057 | 0.999 | 0.409 | 0.068 |
| C3 | 0.120 | 0.924 | 0.357 | 0.188 |
| C4 | 0.035 | 0.999 | 0.159 | 0.737 |
| Normalized disease severity | | | | |
|  | **CSA (mm²)** | | **Eccentricity** | |
|  | **r** | **p-value** | **r** | **p-value** |
| C1 | -0.217 | 0.380 | 0.012 | 0.999 |
| C2 | -0.248 | 0.267 | 0.131 | 0.863 |
| C3 | -0.157 | 0.679 | 0.113 | 0.990 |
| C4 | -0.030 | 0.999 | -0.111 | 0.999 |
