## Supplementary Table5 for "Spinal cord damage in Friedreich’s ataxia: Results from the ENIGMA-Ataxia"

**Supplementary Table5:** Results of ROI-based analyses to assess CSA damage at each stage of FRDA (DD1: Time from ataxia onset <5 years, DD2: Time from ataxia onset between 5-10 years, DD3: Time from ataxia onset between 10-15 years, DD4: Time from ataxia onset between 15-20 years, DD5: Time from ataxia onset > 20 years).

| Veterbral Labels | Subgroup | Measures | Controls (mean±SD) | FRDA (mean±SD) | Bonferroni | Effect Size |
| --- | --- | --- | --- | --- | --- | --- |
| C1 | DD1 | Area (mm²) | 64.2 ± 7.1 | 48.0 ± 8.0 | <0.001 | 2.12 |
|  |  | ECC | 0.717 ± 0.057 | 0.782 ± 0.042 | <0.001 | 1.30 |
|  | DD2 | Area (mm²) | 66.3 ± 8.1 | 46.3 ± 8.8 | <0.001 | 2.36 |
|  |  | ECC | 0.709 ± 0.073 | 0.800 ± 0.052 | <0.001 | 1.46 |
|  | DD3 | Area (mm²) | 64.3 ± 7.1 | 42.5 ± 6.9 | <0.001 | 3.12 |
|  |  | ECC | 0.713 ± 0.076 | 0.779 ± 0.057 | <0.001 | 0.98 |
|  | DD4 | Area (mm²) | 64.8 ± 5.7 | 41.1 ± 7.5 | <0.001 | 3.54 |
|  |  | ECC | 0.700 ± 0.061 | 0.782 ± 0.051 | <0.001 | 1.46 |
|  | DD5 | Area (mm²) | 63.5 ± 7.3 | 38.8 ± 7.9 | <0.001 | 3.23 |
|  |  | ECC | 0.702 ± 0.071 | 0.764 ± 0.059 | <0.001 | 0.96 |
| C2 | DD1 | Area (mm²) | 63.2 ± 7.6 | 46.9 ± 6.7 | <0.001 | 2.31 |
|  |  | ECC | 0.759 ± 0.047 | 0.814 ± 0.029 | <0.001 | 1.44 |
|  | DD2 | Area (mm²) | 64.1 ± 7.2 | 45.0 ± 7.8 | <0.001 | 2.53 |
|  |  | ECC | 0.755 ± 0.044 | 0.825± 0.040 | <0.001 | 1.66 |
|  | DD3 | Area (mm²) | 62.9 ± 7.1 | 41.4 ± 6.9 | <0.001 | 3.08 |
|  |  | ECC | 0.755 ± 0.055 | 0.820 ± 0.036 | <0.001 | 1.41 |
|  | DD4 | Area (mm²) | 63.1 ± 5.9 | 40.5 ± 7.5 | <0.001 | 3.32 |
|  |  | ECC | 0.727 ± 0.055 | 0.808 ± 0.048 | <0.001 | 1.57 |
|  | DD5 | Area (mm²) | 61.7 ± 7.1 | 38.8 ± 8.7 | <0.001 | 2.86 |
|  |  | ECC | 0.749 ± 0.061 | 0.804 ± 0.049 | <0.001 | 1.01 |
| C3 | DD1 | Area (mm²) | 63.1 ± 7.0 | 48.0 ± 7.1 | <0.001 | 2.14 |
|  |  | ECC | 0.797 ± 0.041 | 0.844 ± 0.028 | <0.001 | 1.37 |
|  | DD2 | Area (mm²) | 64.1 ± 7.4 | 46.0 ± 8.0 | <0.001 | 2.35 |
|  |  | ECC | 0.787 ± 0.046 | 0.852 ± 0.036 | <0.001 | 1.60 |
|  | DD3 | Area (mm²) | 62.7 ± 7.4 | 42.0 ± 8.1 | <0.001 | 2.65 |
|  |  | ECC | 0.784 ± 0.047 | 0.844 ± 0.034 | <0.001 | 1.47 |
|  | DD4 | Area (mm²) | 62.2 ± 8.0 | 41.5 ± 8.6 | <0.001 | 2.48 |
|  |  | ECC | 0.763 ± 0.059 | 0.833 ± 0.044 | <0.001 | 1.33 |
|  | DD5 | Area (mm²) | 61.5 ± 7.6 | 39.4 ± 8.1 | <0.001 | 2.80 |
|  |  | ECC | 0.796 ± 0.049 | 0.835 ± 0.045 | <0.001 | 0.83 |
| C4 | DD1 | Area (mm²) | 65.1 ± 6.5 | 50.6 ± 8.8 | <0.001 | 1.83 |
|  |  | ECC | 0.826 ± 0.040 | 0.871 ± 0.024 | <0.001 | 1.39 |
|  | DD2 | Area (mm²) | 66.0 ± 7.7 | 47.1 ± 8.8 | <0.001 | 2.28 |
|  |  | ECC | 0.820 ± 0.042 | 0.874 ± 0.030 | <0.001 | 1.52 |
|  | DD3 | Area (mm²) | 63.9 ± 7.7 | 45.3 ± 7.6 | <0.001 | 2.42 |
|  |  | ECC | 0.818 ± 0.033 | 0.868 ± 0.027 | <0.001 | 1.69 |
|  | DD4 | Area (mm²) | 62.5 ± 8.3 | 42.7 ± 9.2 | <0.001 | 2.27 |
|  |  | ECC | 0.813 ± 0.036 | 0.856 ± 0.040 | 0.003 | 1.16 |
|  | DD5 | Area (mm²) | 63.3 ± 7.4 | 39.8 ± 8.3 | <0.001 | 3.00 |
|  |  | ECC | 0.829 ± 0.037 | 0.868 ± 0.032 | <0.001 | 1.11 |
