## Supplementary Table6 for "Spinal cord damage in Friedreich’s ataxia: Results from the ENIGMA-Ataxia"

**Supplementary Table6:** Results of ROI-based analyses to assess CSA damage at each stage of FRDA (DS1: Normalized disease severity <0.25, DS2: Normalized disease severity between 0.26-0.50, DS3: Normalized disease severity between 0.51-0.75, DS4: Normalized disease severity >0.75). All comparisons are significant, i.e., p<0.001 after Bonferroni correction.

| Veterbral Labels | Subgroup | Structures | Controls (mean±SD) | FRDA (mean±SD) | Bonferroni | Effect Size |
| --- | --- | --- | --- | --- | --- | --- |
| C1 | DS1 | Area (mm²) | 68.7 ± 7.6 | 48.7 ± 8.7 | <0.001 | 2.49 |
|  |  | ECC | 0.704 ± 0.072 | 0.793 ± 0.054 | <0.001 | 1.42 |
|  | DS2 | Area (mm²) | 64.9 ± 7.4 | 45.5 ± 7.9 | <0.001 | 2.99 |
|  |  | ECC | 0.714 ± 0.068 | 0.784 ± 0.056 | <0.001 | 1.13 |
|  | DS3 | Area (mm²) | 64.1 ± 6.4 | 39.3± 7.0 | <0.001 | 3.78 |
|  |  | ECC | 0.702 ± 0.068 | 0.772 ± 0.054 | <0.001 | 1.17 |
|  | DS4 | Area (mm²) | 62.0 ± 6.1 | 36.9 ± 7.2 | <0.001 | 3.64 |
|  |  | ECC | 0.695 ± 0.063 | 0.781± 0.052 | 0.001 | 1.42 |
| C2 | DS1 | Area (mm²) | 66.0 ± 7.0 | 46.7 ± 7.9 | <0.001 | 2.65 |
|  |  | ECC | 0.755 ± 0.047 | 0.825 ± 0.037 | <0.001 | 1.63 |
|  | DS2 | Area (mm²) | 63.3 ± 6.8 | 44.6 ± 7.2 | <0.001 | 3.23 |
|  |  | ECC | 0.756 ± 0.051 | 0.816 ± 0.037 | <0.001 | 1.37 |
|  | DS3 | Area (mm²) | 62.4 ± 6.4 | 38.8 ± 7.6 | <0.001 | 3.39 |
|  |  | ECC | 0.744 ± 0.057 | 0.807 ± 0.049 | <0.001 | 1.21 |
|  | DS4 | Area (mm²) | 62.3 ± 7.1 | 37.0 ± 7.7 | <0.001 | 3.73 |
|  |  | ECC | 0.730± 0.058 | 0.821 ± 0.047 | <0.001 | 1.64 |
| C3 | DS1 | Area (mm²) | 65.2 ± 7.7 | 46.7 ± 9.0 | <0.001 | 2.21 |
|  |  | ECC | 0.802 ± 0.042 | 0.855 ± 0.031 | <0.001 | 1.53 |
|  | DS2 | Area (mm²) | 63.3 ± 7.1 | 46.0 ± 7.6 | <0.001 | 2.74 |
|  |  | ECC | 0.790 ± 0.043 | 0.843 ± 0.035 | <0.001 | 1.37 |
|  | DS3 | Area (mm²) | 61.6 ± 7.1 | 39.5 ± 7.5 | <0.001 | 3.06 |
|  |  | ECC | 0.783 ± 0.051 | 0.834 ± 0.043 | <0.001 | 1.09 |
|  | DS4 | Area (mm²) | 62.8 ± 9.2 | 36.8 ± 7.5 | <0.001 | 3.70 |
|  |  | ECC | 0.778 ± 0.059 | 0.853 ± 0.045 | 0.003 | 1.33 |
| C4 | DS1 | Area (mm²) | 67.0 ± 7.0 | 50.2 ± 8.5 | <0.001 | 2.24 |
|  |  | ECC | 0.823 ± 0.043 | 0.881 ± 0.025 | <0.001 | 1.89 |
|  | DS2 | Area (mm²) | 65.2 ± 7.5 | 47.5 ± 8.7 | <0.001 | 2.52 |
|  |  | ECC | 0.821 ± 0.037 | 0.868 ± 0.028 | <0.001 | 1.54 |
|  | DS3 | Area (mm²) | 63.1 ± 7.2 | 40.2 ± 7.3 | <0.001 | 3.13 |
|  |  | ECC | 0.823 ± 0.039 | 0.864 ± 0.033 | <0.001 | 1.18 |
|  | DS4 | Area (mm²) | 64.4 ± 8.9 | 38.0 ± 8.5 | <0.001 | 2.89 |
|  |  | ECC | 0.834 ± 0.036 | 0.879 ± 0.036 | 0.162 | 0.97 |
